# High Precision ECG Digitization Using Artificial Intelligence

**DOI:** 10.1101/2024.08.31.24312876

**Authors:** Anthony Demolder, Viera Kresnakova, Michal Hojcka, Vladimir Boza, Andrej Iring, Adam Rafajdus, Simon Rovder, Timotej Palus, Martin Herman, Felix Bauer, Viktor Jurasek, Robert Hatala, Jozef Bartunek, Boris Vavrik, Robert Herman

**Author notes:** Co-primary and corresponding authors: Viera Kresnakova.

## Abstract

**Background:** The digitization of electrocardiograms (ECGs) is an important process in modern healthcare, enabling the preservation, transmission, and advanced analysis of ECG data. Traditional methods for digitizing ECGs from paper formats face significant challenges, particularly in real-world scenarios with varying image quality, paper distortions, and overlapping signals. Existing solutions often require manual input and are limited by their dependence on high-quality images and standardized layouts.

**Methods:** This study introduces a fully automated, deep learning-based approach for high-precision ECG digitization, implementing a two-stage process. In the ECG normalization phase, image distortions are corrected, axes are calibrated, and a standardized grid structure is generated. The ECG reconstruction phase uses deep learning techniques to extract and digitize the leads, with subsequent post-processing to refine the digital signal. The tool was evaluated using a custom-built PMcardio ECG Image Database (PM-ECG-ID) comprising 6,000 ECG images generated from 100 unique ECGs, subjected to various augmentations to simulate real-world challenges. Performance was assessed using Pearson’s correlation coefficient (PCC), root mean squared error (RMSE), and signal-to-noise ratio (SNR).

**Results:** The digitization tool demonstrated an average PCC consistently exceeding 91% across all leads, SNR above 12.5 dB and an RMSE below 0.10 mV. The time to ECG digitization was consistently less than 10 seconds. The failure rate was low, averaging 6.62%, with most failures occurring under extreme conditions such as severe blurring or significant image degradation. The tool maintained robust performance even under challenging scenarios, such as low-resolution images, distorted grids, and overlapping signals. Performance metrics indicated slight variations across different leads, with leads V3-V5 showing marginally higher RMSE and lower PCC, reflecting the complexity of digitizing large amplitude signals.

**Conclusion:** Our deep learning-based approach for ECG digitization delivers high precision and reliability, effectively addressing real-world challenges such as image distortions, lighting variations, and overlapping signals. This fully automated method enhances the accessibility and utility of ECG data, supporting advanced AI-driven analysis in cardiac healthcare.

## Introduction

The electrocardiogram (ECG) is a widely accessible diagnostic tool used to evaluate the electrical activity of the heart. Despite its essential role in diagnosing and monitoring cardiac abnormalities, many healthcare settings still rely on paper-based ECGs. Moreover, ECGs are performed at every healthcare level and their transmission throughout the patient continuum is hindered by the diverse range of device manufacturers. Therefore, these systems depend on the transmission of paper copies, which are susceptible to quality deterioration, including ink fading, blurring, folding, stains, or on ECG images that may contain patient-identifiable information and do not meet compliance standards. In contrast, digital ECG files can be stored indefinitely while retaining high quality, enabling advanced ECG analysis using modern artificial intelligence algorithms^1–6^ and easy, compliant transmission between healthcare professionals.

Several approaches for converting ECG images into digital waveforms (ECG digitization) have been proposed over the past decades. Initially, ECGScan^7^ offered extensive image file compatibility, grid detection, and waveform reconstruction, but was limited by significant user input. The fully automated ECG digitization tool from Imperial College London^8^ utilized deep learning to convert paper ECGs into digital signals. However, the tool struggles with lead labels detection in low-resolution images and overlapping signals, which may affect its applicability across different ECG abnormalities. Recently, ECGMiner^9^, an open-source solution, showed high accuracy in the automated digitization of ECG images. However, it faced challenges such as interpreting lead labels, had issues with overlapping leads, and depended on high-quality image inputs. Moreover, most existing approaches require manual user input, are limited to high-quality scans, and fail to cover the complex, real-world spectrum of ECG layouts, lighting conditions, lead visibility, or overlapping waveforms (Table 1)^7–10^.

**Table 1.** Comparison of ECG Processing Models. **Comprehensive normalization** includes handling distortions, bends, crumples, and shadows across various real-world scenarios, without a significant drop in performance metrics. **Handling leads overlapping** refers to the ability to accurately digitize ECG signals when leads overlay, without significant performance drops. **Automatic format and rotation detection** is based solely on the orientation and layout of the leads. **Rapid digitization** is defined as completing the digitization process in under 10 seconds. The dash (’–’) suggests that it is unclear or not specified if the feature is supported by the algorithm.

|  | Our<br>solution | Compared solutions |  |  |  |  |  |  |  |
| --- | --- | --- | --- | --- | --- | --- | --- | --- | --- |
|  |  | Santamónica<sup>9</sup> | Wu<sup>8</sup> | Badilini<sup>7</sup> | Mishra<sup>21</sup> | Isabel<sup>22</sup> | Ravichandran<sup>23</sup> | Baydoun<sup>24</sup> | Fortune<sup>25</sup> |
| Fully automated | ✓ | ✓ | ✓ | – | – | – | – | – | ✓ |
| Comprehensive normalization | ✓ | – | – | – | – | – | – | – | – |
| Handling leads overlapping | ✓ | – | – | – | – | – | – | – | – |
| Automatic format detection | ✓ | – | – | – | – | – | – | – | – |
| Automatic rotation detection | ✓ | – | – | – | – | – | – | – | – |
| Rapid digitization | ✓ | ✓ | – | – | – | – | – | – | ✓ |

There is no single method that reliably and automatically generates a digital waveform from any image of a 12-lead ECG, regardless of input quality, manufacturer, or environment where the image is captured. We aimed to develop a solution for ECG digitization that is smartphone-based, versatile, and fully automated end-to-end. We evaluated our approach using a large, independent database of ECG images spanning various cardiac abnormalities captured in scenarios simulating real-world challenges for ECG digitization.

## Methods

### Aims and supported settings

The goal of our ECG digitization solution is to convert image inputs (e.g., photographs of ECG printouts or monitors) into digital waveforms. Our ECG digitization tool operates under certain requirements to ensure coverage of various ECG formats. A table of supported ECG machine settings and real-world scenarios is available in the supplements (Supplemental Table 1).

The ECG configuration specifies the number of rows, columns, and rhythm leads in the following format: [Rows] + [Columns] + [Rhythm leads]. The lead layout determines the order of the leads to be supported, with the possibility that a single ECG may span multiple pages. Supplemental Table 2 compares the supported formats and highlights the limitations of existing ECG digitization tools, which are often restricted to specific configurations and lack automated detection of the ECG format.

### Digitization tool

The ECG digitization tool (Figure 1) consists of two key stages, **ECG Normalization** and **ECG Reconstruction**. In the initial stage, images are standardized based on the underlying millimeter grid, resolving any distortions caused by paper bends, crumples, or variations in image perspective. In the second stage, waveforms are segmented into masks and subsequently classified into individual leads using ECG format detection. An example output of the digitization platform is available in Figure 2.

**Figure 1.**
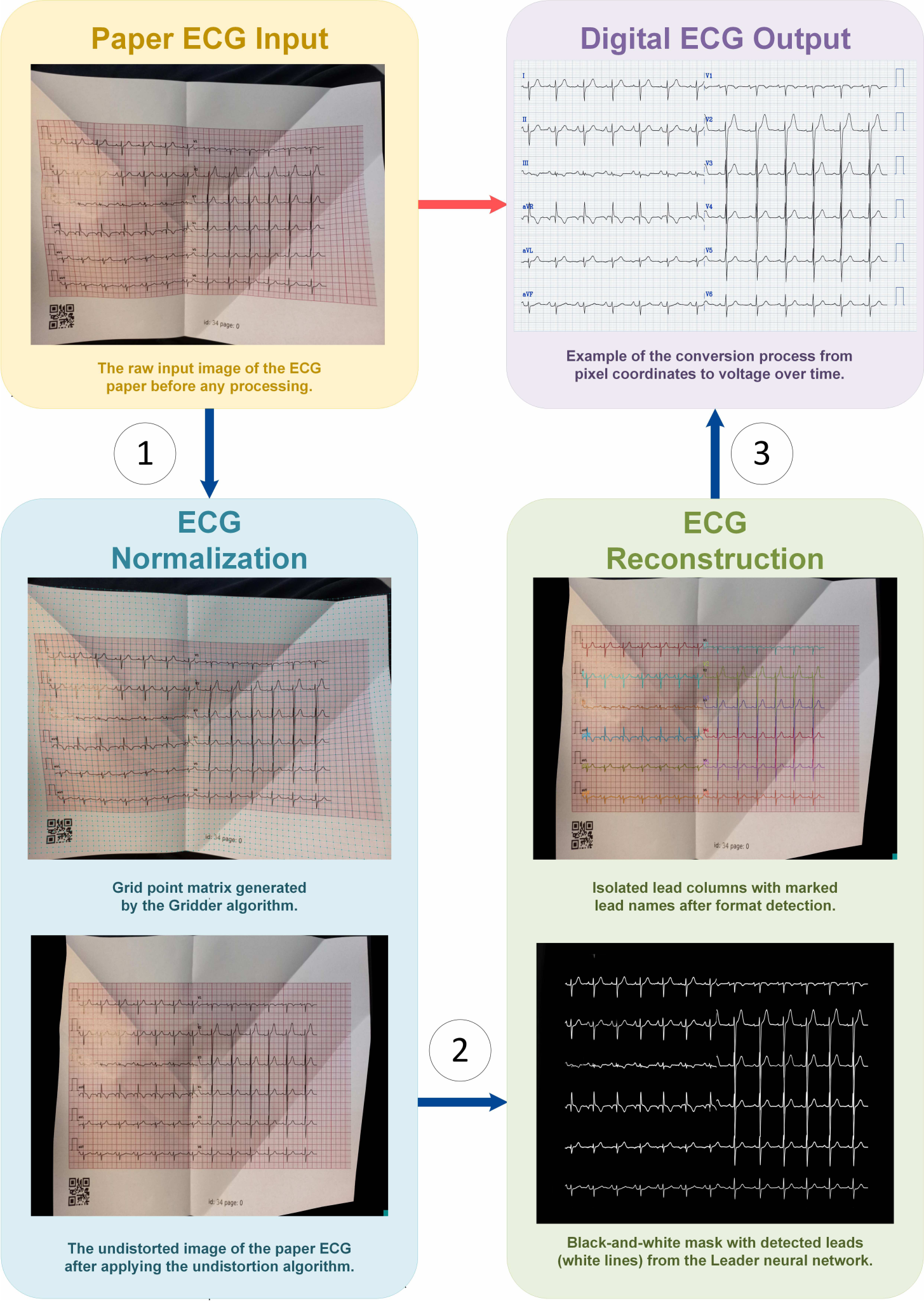
The process flow of converting paper ECG input into digital ECG output, illustrating the steps of ECG normalization and ECG reconstruction.

**Figure 2.**
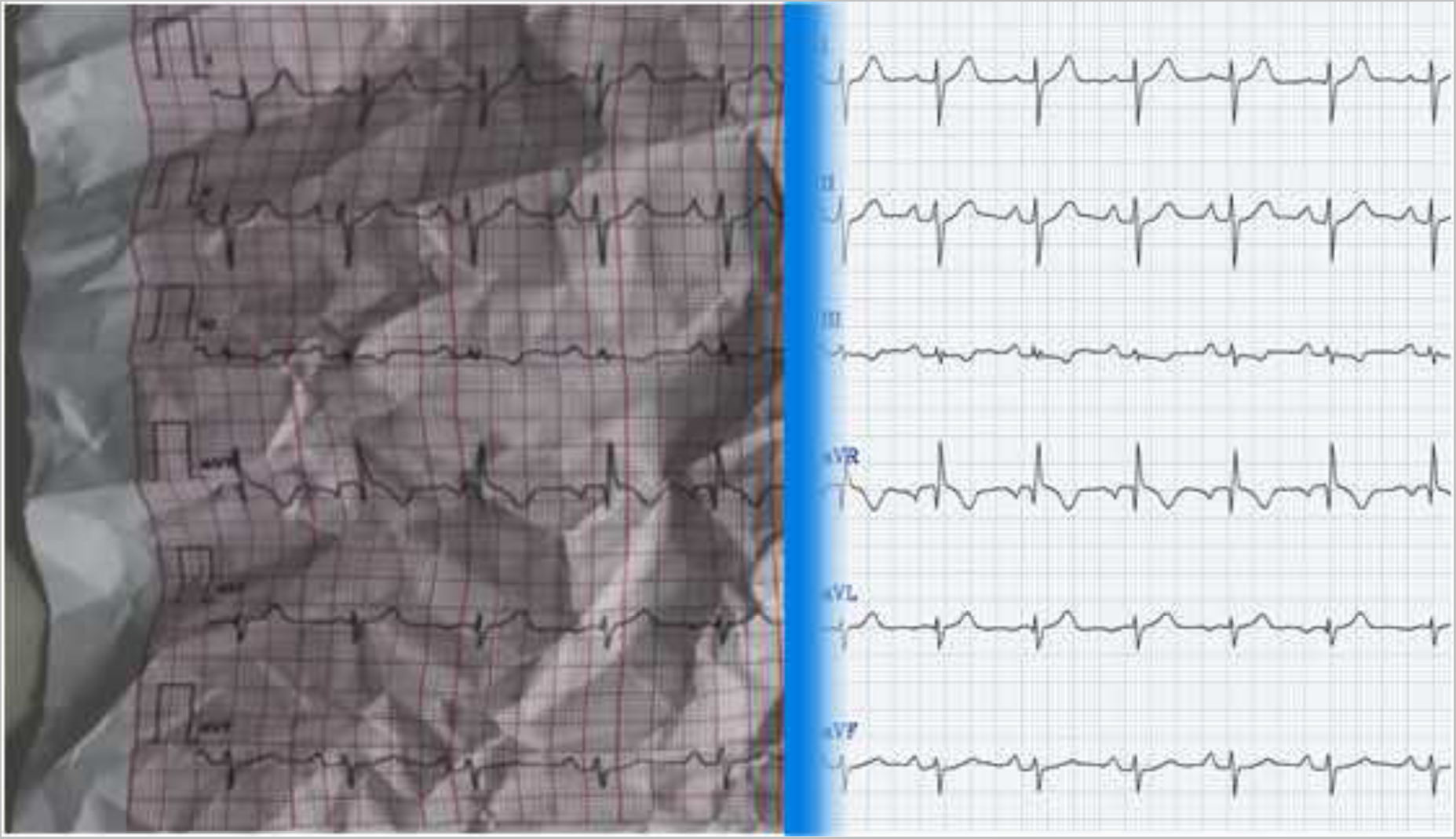
Comparison of a crumpled paper ECG with a digitization version: highlighting the difference in clarity and readability

### ECG Normalization

#### Data sources and pre-processing

The goal of ECG normalization is to identify all major grid intersections on standard 12-lead ECG images. A total of 176 ECG images were annotated, with an average of 1944 (±649) grid intersections marked per ECG. The dataset was randomly split into 140 training images and 36 validation images. The annotated ECG images displayed several variations: extreme camera positions, different paper qualities (bends, distortions, scratches), various visual effects (brightness, contrast, blurriness, color tones, shadows, Moiré effect), and various ECG printout formats (lead layouts, grid shapes, ink colors, paper sizes). To enhance robustness, over 10,000 augmented input patches were created from the original images, covering a range of visual conditions. The input patches, each measuring 256 ⇥ 256 pixels, were obtained by segmenting the ECG images into smaller sections. Figure 3 illustrates the various data augmentations applied to these ECG patches.

**Figure 3.**
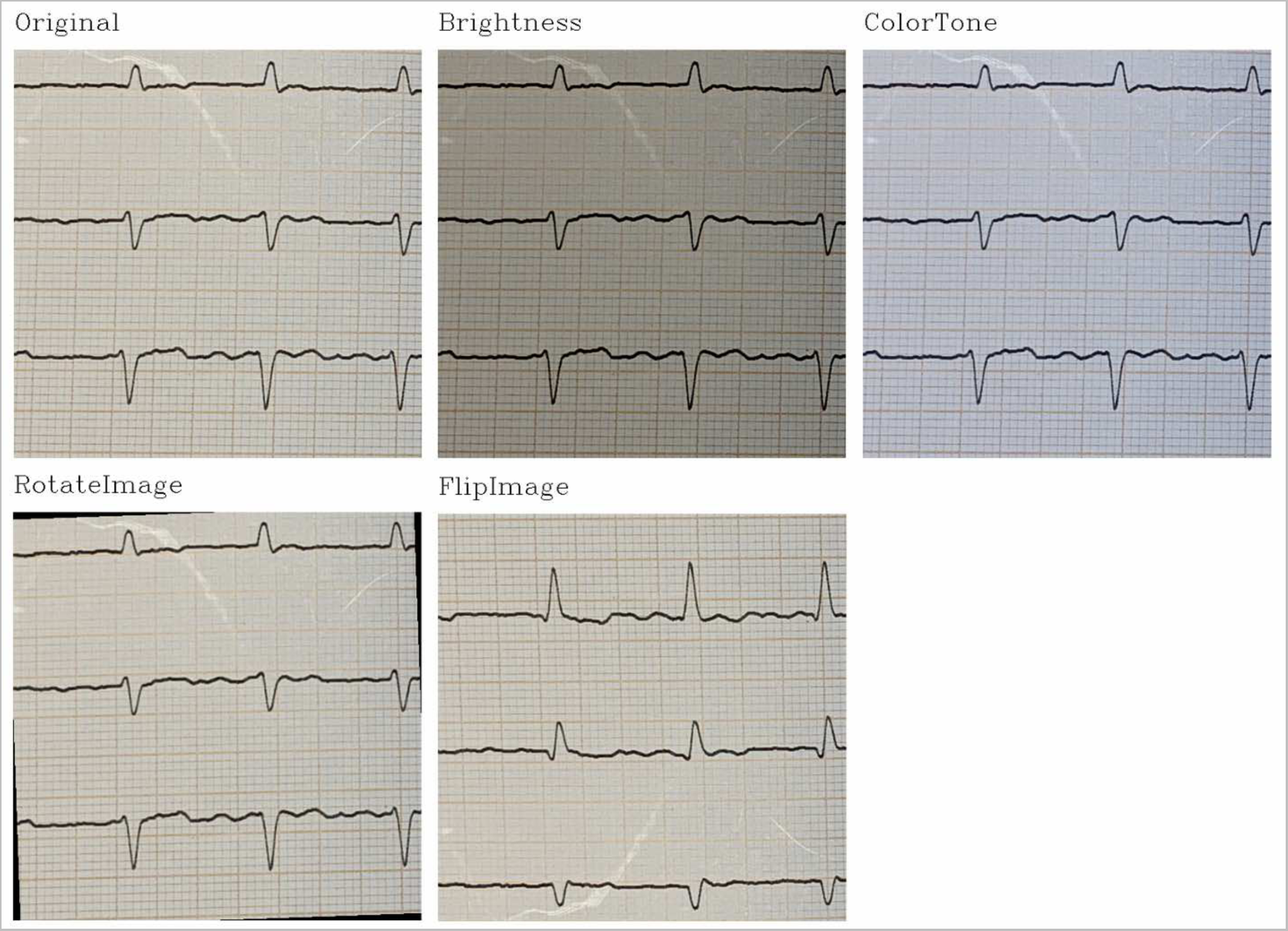
Example of different types of data augmentations are applied to ECG patches. From left to right: brightness adjustment, color tone variation, rotation, and image flipping.

#### Deep learning model configuration

Our patent-pending approach utilizes the U-net architecture integrated with Residual Blocks (ResBlocks) to create a model that outputs a segmented mask of grid intersections^11–13^. The model was trained for 300 epochs with a learning rate of 0.005 using ADAM optimizer, binary cross-entropy with logits loss, and the SiLU activation function for non-linearity^14,15^. The output of the neural network is a black-and-white mask with the identified dots, which we refer to as Dotter.

#### Post-processing of the normalization outputs

Post-processing includes two steps: the Gridder algorithm and the Undistortion algorithm. The Gridder algorithm constructs a grid matrix from Dotter’s output, handling distortions and filling gaps through interpolation. The finalized grid, derived from the Gridder algorithm, is visualized in Figure 1. The Undistortion algorithm corrects nonlinear paper distortions by applying a 4-point linear undistortion method to each grid square individually, effectively reconstructing the undistorted image (see Figure 4). The process requires an ECG photo and precise gridline intersection coordinates. Further details can be found in the patent document by Rovder et al^16^.

**Figure 4.**
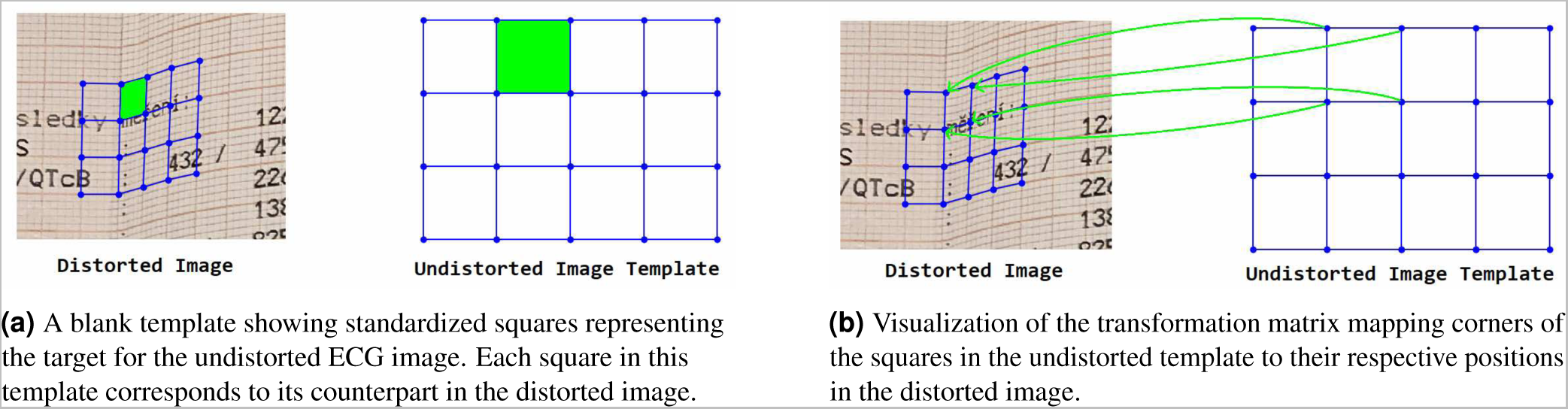
Various representations of the undistorted template and its transformation mapping.

### ECG Reconstruction

The reconstruction process involves two key steps: detecting all waveforms in the image using deep neural networks and classifying them into individual ECG leads through heuristic post-processing.

### Data sources and pre-processing

The ECG reconstruction requires annotations of the leads rather than grid intersections. A total of 232 ECG images were used, with 185 designated for training and 47 for validation. Annotators were instructed to open ECG images and create a layer for each visible lead. Each label was verified by plotting the layers over the original photo and visually checking to ensure precise segmentation of all leads. Over 10,000 input patches were generated from the training set to improve model robustness, applying the same data augmentations as those used for the Dotter neural network.

### Deep learning model configuration

The model for ECG reconstruction was a U-Net with ResBlocks. Accuracy was measured using the Intersection over Union (IoU), also known as the Jaccard index, which quantifies the overlap between predicted and actual segments. The output of the Leader neural network is a black-and-white mask, as shown in the ECG Reconstruction part of Figure 1, with the detected leads represented by white lines.

### Post-processing of the reconstruction outputs

Post-processing refines the detected leads into digital signals by filtering noise, identifying the region of interest (ROI), and segmenting lead columns. The algorithm identifies the baseline for each lead and extends detected paths. In complex regions, it connects lead endpoints, maximizes brightness, and corrects for overlapping leads or gaps to improve accuracy.

### Automatic Format Detection

In the next step, referred to as Format Detection, the optimal photo rotation, ECG layout, and lead order (standard or Cabrera) are determined. This information can be provided as input by the user or determined automatically (currently not possible for multi-photo ECGs) based on the limb-lead logic and rhythm lead information. The final step converts pixel coordinates into ECG signals, requiring user inputs for paper speed and voltage gain to ensure accurate voltage representation. An example of this conversion process is shown as Digital ECG output in Figure 1.

### PM-ECG-ID database

The PM-ECG-ID database was created to offer a diverse and comprehensive resource for the evaluation and benchmarking of ECG digitization approaches. Our database aims to provide wide coverage of real-world scenarios, enabling researchers to identify limitations and improve performance. The ECGs in the PM-ECG-ID database were randomly sampled from the publicly available PTB-XL dataset^17,18^. A total of 75 ECGs were systematically chosen using the available diagnostic statements to reflect 15 commonly prevalent diagnostic patterns, including arrhythmias, infarctions, or conduction abnormalities. In addition, 25 sinus rhythm ECGs were added to the dataset. The base dataset comprised 100 unique standard 12-lead ECGs (some multipage, resulting in 110 visualizations), recorded at a sampling frequency of 500 Hz.

### ECG printout variations and image augmentations

ECG waveforms were visualized on a millimeter grid, printed, and then photographed under various scenarios, including:

- paper transformations (such as paper crumples or bends),
- image quality modifications (contrast, brightness, blur, or screen glare)
- image processing (compression, resolution)
- distances (large, medium, small)
- phones (Samsung, iPhone or Doogee)

In addition to ECG printouts, additional augmentations were performed using the base ECG waveforms before printing. A full schematic overview of the various ECG image variations and their creation is available in Figure 5.

**Figure 5.**
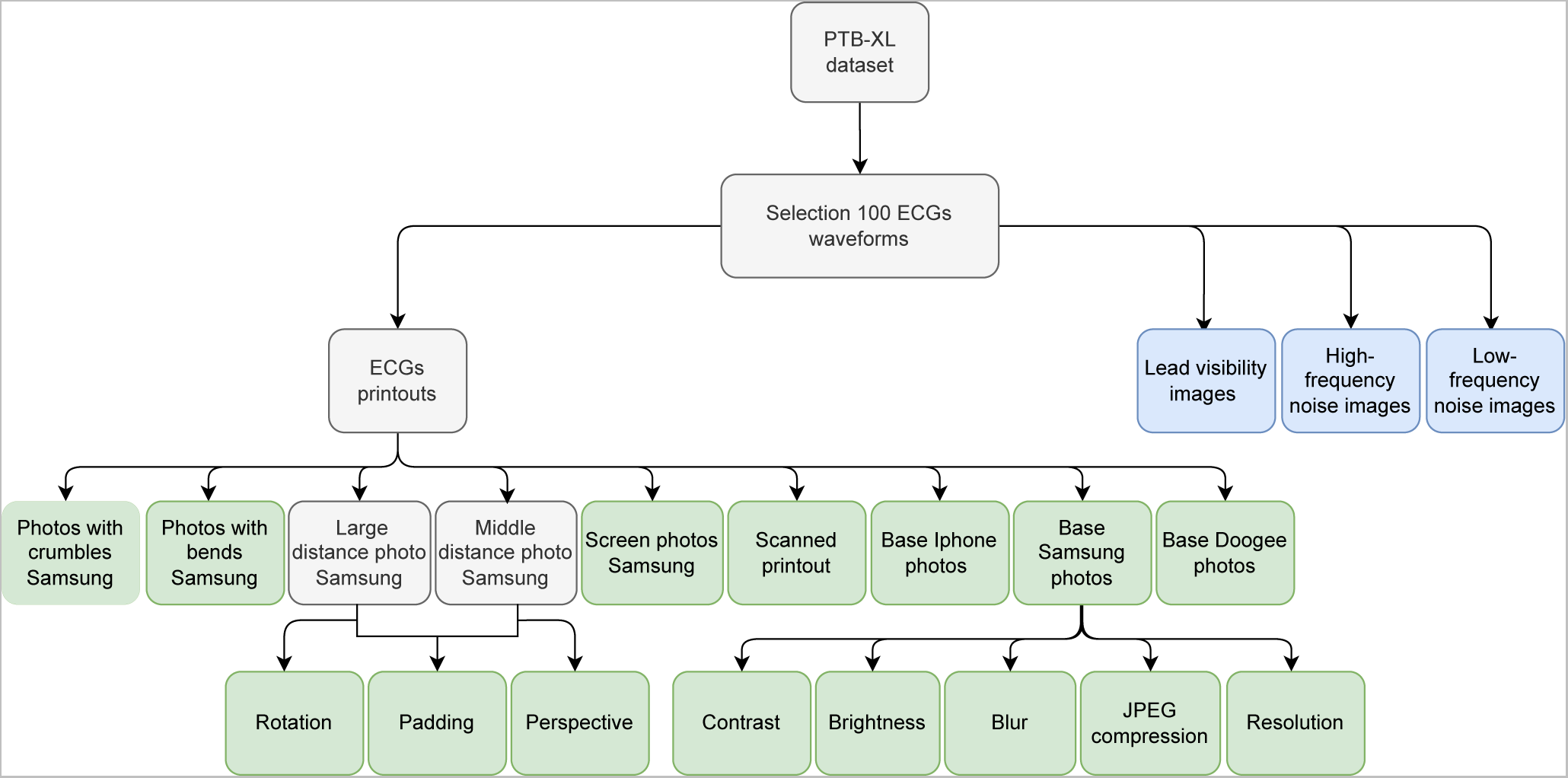
Schematic overview of the creation of the ECG digitization database. The blue sections depict the final augmentation of ECG waveforms, saved in JPEG format. The green sections represent variations and augmentations created on printouts of ECGs.

The final PM-ECG-ID database consists of 6,000 unique ECG images across 60 real-world scenarios. More information regarding the distribution of ECG images across various image scenarios is available in Table 1 (Appendix 1) and multiple scenarios are illustrated in Figure 6. The data records are publicly accessible and stored in the Figshare repository: https://doi.org/10.5281/zenodo.1361767219.

**Figure 6.**
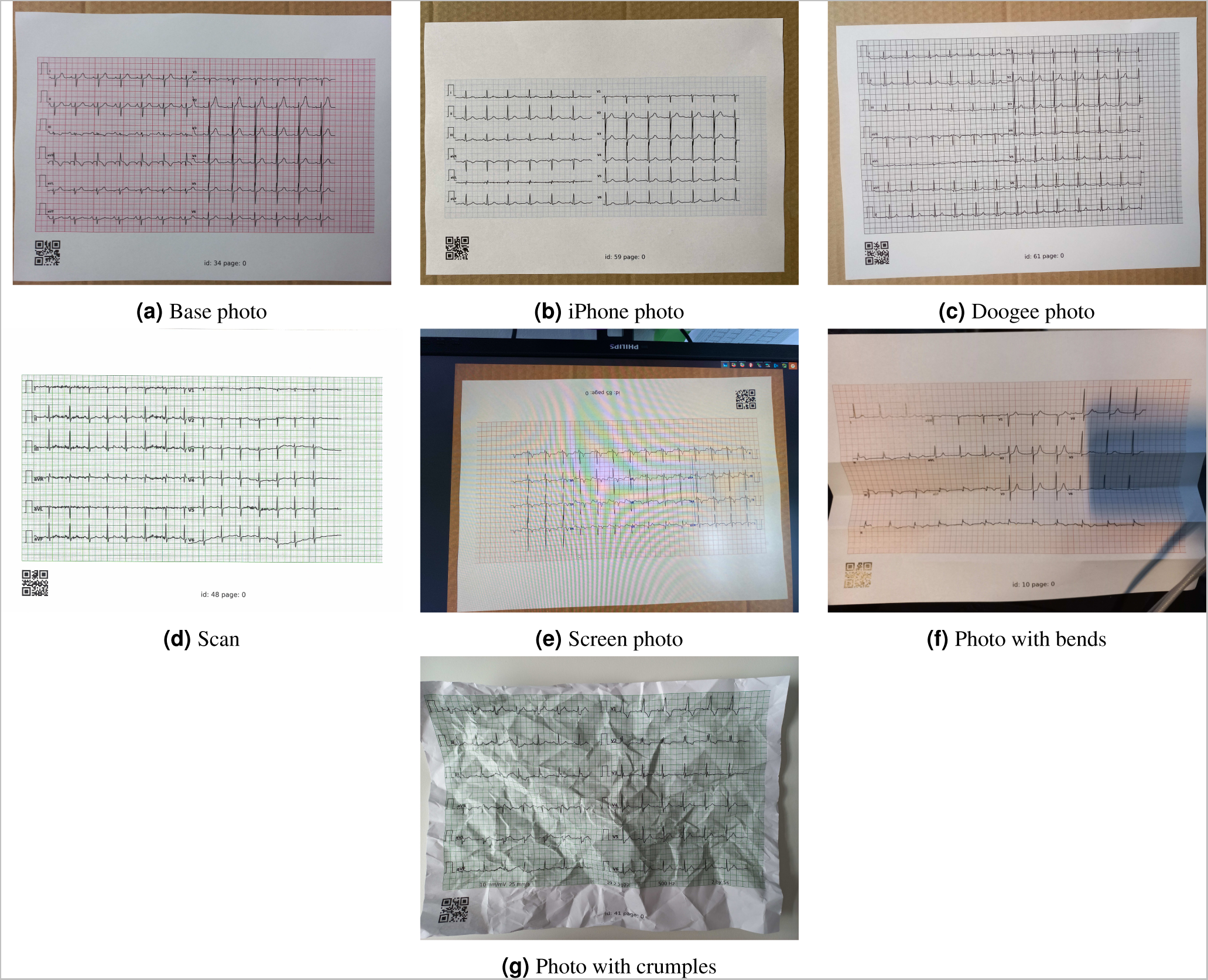
Example of different variations of photos of ECG printouts from the PM-ECG-ID.

### Statistical analysis

To compare digitized and original ECG signals, a temporal alignment procedure adjusted the digitized signals to match the original leads within a 0.5-second shift range. Padding with NaNs and evaluating coverage identified the optimal shift, while a vertical shift correction minimized offsets by subtracting the median difference between signals. These adjustments reduced discrepancies, ensuring accurate metrics. The PM-ECG-ID Database was used for validation. The association between digitized ECG signals and original ECG signals was evaluated using Pearson’s correlation coefficient (PCC), root mean squared error (RMSE), and the signal-to-noise ratio (SNR). A detailed description of each metric used can be found in Supplemental Table 3. An analysis was conducted to evaluate the performance of ECG digitization across different leads and real-world image scenarios. Time to ECG digitization was calculated and compared across scenarios. Reported values are described as mean (± standard deviation) unless otherwise specified. A P-value of <0.01 was considered statistically significant.

## Results

### Performance across leads

Our ECG digitization tool performance across three metrics on an independent validation set of 6,000 real-world scenario ECG images is presented in Table 2. The average PCC consistently exceeded 91% across all leads, and the average RMSE remained below 0.10 mV in all successfully digitized cases. The PCC and RMSE showed slight variations across different leads (P<0.001). Lower performance was recorded in leads V3-V5, which tend to display the largest signal amplitudes, with a lower PCC, higher RMSE, and lower SNR, respectively (P<0.001 for each). The average digitization fail rate was 6.62% and did not differ across leads (*P* ⇡ 1). To better understand the overall impact of digitization failures, metrics were also calculated for the entire database, with failures treated as having an RMSE equivalent to the RMSE of an all-zero signal, with PCC and SNR set to zero (Supplemental Table 4 and 5).

**Table 2.** Summary of lead-specific and overall performance metrics for 6,000 ECG entries (100 unique ECGs with 60 augmentations per ECG). The table evaluates four key parameters: average fail rate (%), RMSE, Pearson Correlation Coefficient (PCC), and Signal-to-Noise Ratio (SNR).

| Lead | Average<br>fail rate | RMSE | PCC | SNR |
| --- | --- | --- | --- | --- |
| I | 6.63% | 0.034 | 0.953 | 15.574 |
| II | 6.65% | 0.040 | 0.957 | 15.717 |
| III | 6.65% | 0.043 | 0.957 | 15.762 |
| aVR | 6.62% | 0.031 | 0.949 | 15.232 |
| aVL | 6.62% | 0.034 | 0.955 | 15.446 |
| aVF | 6.62% | 0.035 | 0.958 | 15.758 |
| V1 | 6.57% | 0.045 | 0.957 | 16.445 |
| V2 | 6.57% | 0.079 | 0.949 | 15.117 |
| V3 | 6.57% | 0.095 | 0.936 | 13.799 |
| V4 | 6.67% | 0.099 | 0.918 | 12.758 |
| V5 | 6.67% | 0.081 | 0.935 | 13.484 |
| V6 | 6.65% | 0.051 | 0.952 | 15.505 |
| All leads | 6.62% | 0.056 | 0.948 | 15.050 |

### Performance across image scenarios

The quality of ECG digitization across various real-world conditions is presented in Table 3, with image subsets empirically classified into easy, medium, or hard categories of complexity based on the input quality, extent of image distortions, noise levels, and overall signal visibility. In the easy category group (12 subsets), the average PCC was 0.977 (0.975 to 0.981), RMSE was 0.041 mV (0.037 to 0.043 mV), and the average SNR was 16.682 (16.239 to 17.644). In the medium complexity category (29 subsets), which included conditions such as moderate blurring and contrast variations, the average PCC, RMSE, and SNR were 0.970 (0.940 to 0.985), 0.047 mV (0.035 to 0.069 mV), and 15.785 (11.922 to 21.225) respectively. In the hardest complexity group (19 subsets), extreme transformations such as ’Blur kernel 41’ and high degrees of rotation significantly impacted the ECG image input quality. There was a substantial reduction across all three metrics, with the PCC averaging 0.817 (0.087 to 0.968), RMSE 0.098 mV (0.048 to 0.254 mV), and SNR of 11.101 (-0.759 to 22.058). Despite the challenges posed by these extreme conditions, around 80% of the dataset was successfully digitized, albeit with reduced accuracy compared to higher-quality inputs. Failure rates of individual ECG image scenarios indicate instances where digitization quality significantly degraded, primarily attributed to extreme cases or low-quality data inputs. For example, severe scaling ("Scale down 1/32"), extreme blurring ("Blur kernel 31" and "Blur kernel 41"), excessive brightness reduction ("Brightness -160"), or large image capture distance ("Zoom out extreme") showed considerably higher failure rates. Examples of ECG digitization in easy, medium, or hard categories of complexity are illustrated in Figure 7, Figure 8 and Figure 9, respectively.

**Figure 7.**
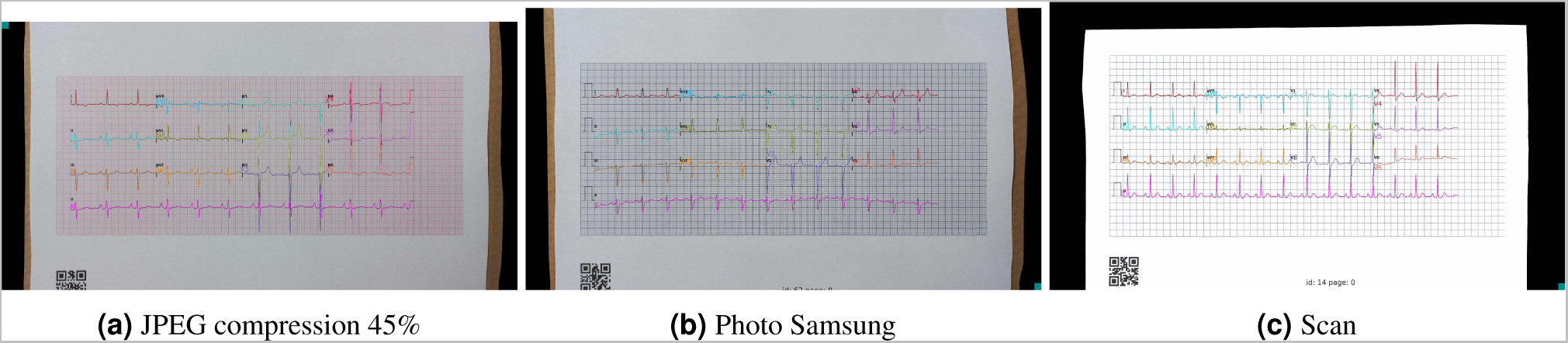
Examples of ECG digitization under high-quality conditions, demonstrating the system’s capability to produce accurate results even with alterations such as JPEG compression at 45%. Each subfigure exemplifies the high fidelity of digitized signals from varied sources, such as compressed images, high-resolution photographs from Samsung devices, and scanned images, highlighting the precision and robustness of the digitization process in optimal scenarios.

**Figure 8.**
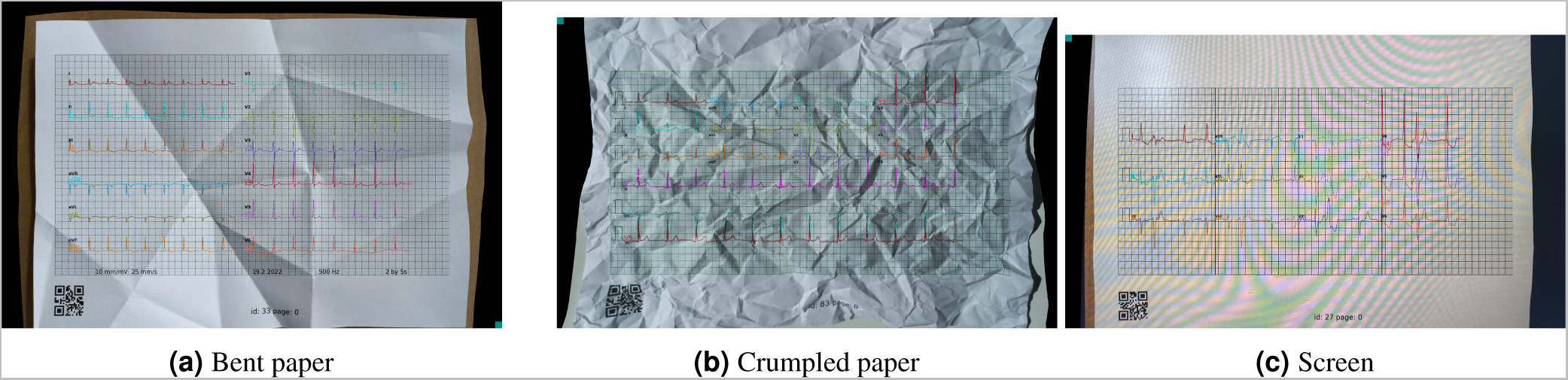
Examples of ECG digitization under medium-quality conditions, showcasing the system’s ability to manage moderate physical distortions and varied display types. Each image illustrates the digitization process’s effectiveness in maintaining data integrity under non-ideal but common operational scenarios.

**Figure 9.**
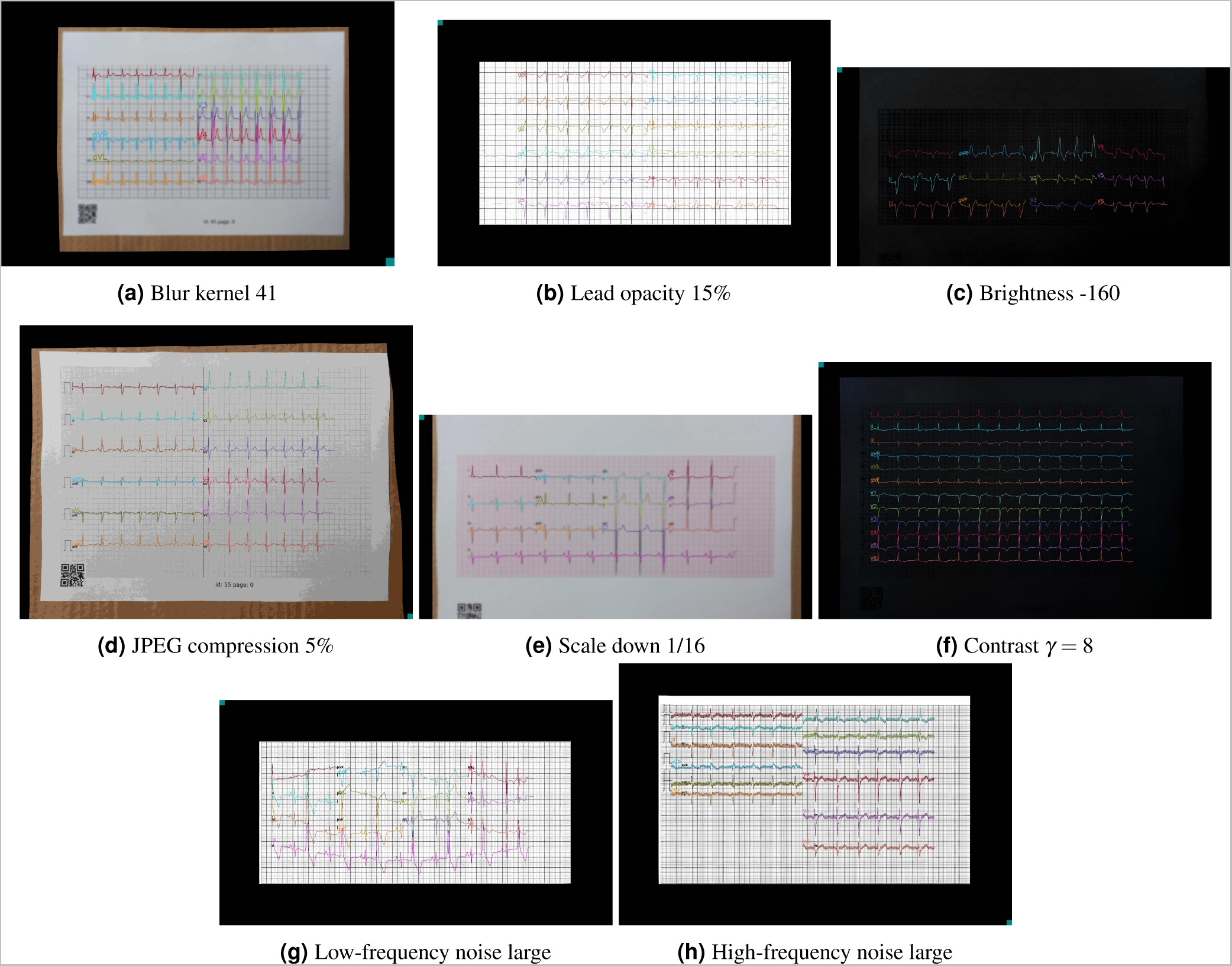
Examples of ECG digitization under challenging conditions. Each subfigure shows the effect of extreme manipulations on the ECG signal quality.

**Table 3.** Comprehensive Evaluation of ECG Digitization Quality Across Varied Conditions: This table details the performance metrics of ECG digitization processes across different datasets subjected to various conditions such as compression, brightness adjustment, and digital noise. Each dataset contains 100 ECGs and is assessed based on Root Mean Square Error (RMSE), Pearson Correlation Coefficient (PCC), and Signal-to-Noise Ratio (SNR).

| Subset | failed | RMSE | PCC | SNR | Average time [s] | Complexity group |
| --- | --- | --- | --- | --- | --- | --- |
| Digital image original | 0.00% | 0.037 | 0.981 | 17.644 | 4.915 | easy |
| JPEG compression 90% | 0.00% | 0.041 | 0.977 | 16.608 | 4.593 | easy |
| JPEG compression 45% | 0.00% | 0.042 | 0.976 | 16.526 | 4.509 | easy |
| JPEG compression 22% | 0.00% | 0.042 | 0.976 | 16.473 | 4.504 | medium |
| JPEG compression 11% | 0.00% | 0.044 | 0.974 | 16.148 | 4.510 | medium |
| JPEG compression 5% | 0.00% | 0.049 | 0.968 | 15.258 | 4.507 | hard |
| JPEG compression 2% | 0.00% | 0.064 | 0.945 | 13.449 | 4.456 | hard |
| Blur kernel 11 | 0.00% | 0.046 | 0.972 | 15.771 | 4.567 | medium |
| Blur kernel 21 | 1.00% | 0.060 | 0.957 | 13.445 | 5.391 | medium |
| Blur kernel 31 | 69.00% | 0.240 | 0.365 | -0.116 | 6.128 | hard |
| Blur kernel 41 | 96.00% | 0.226 | 0.087 | -0.549 | 3.883 | hard |
| Brightness -40 | 0.00% | 0.041 | 0.977 | 16.581 | 4.577 | easy |
| Brightness -80 | 0.00% | 0.043 | 0.974 | 16.292 | 4.539 | medium |
| Brightness -120 | 0.00% | 0.046 | 0.971 | 15.676 | 4.451 | medium |
| Brightness -160 | 5.25% | 0.072 | 0.916 | 12.225 | 5.056 | hard |
| Brightness +40 | 0.00% | 0.041 | 0.977 | 16.669 | 4.525 | easy |
| Brightness +80 | 0.00% | 0.042 | 0.976 | 16.603 | 4.566 | medium |
| Brightness +120 | 0.00% | 0.042 | 0.976 | 16.446 | 4.641 | medium |
| Brightness +160 | 0.00% | 0.048 | 0.963 | 15.617 | 4.679 | hard |
| Contrast $\gamma = 0.125$ | 0.00% | 0.040 | 0.977 | 16.688 | 4.633 | medium |
| Contrast $\gamma = 0.25$ | 0.00% | 0.039 | 0.978 | 16.909 | 4.567 | medium |
| Contrast $\gamma = 0.5$ | 0.00% | 0.040 | 0.977 | 16.823 | 4.537 | easy |
| Contrast $\gamma = 0.666$ | 0.00% | 0.040 | 0.977 | 16.757 | 4.561 | easy |
| Contrast $\gamma = 1.5$ | 0.00% | 0.042 | 0.976 | 16.543 | 4.580 | easy |
| Contrast $\gamma = 2$ | 0.00% | 0.041 | 0.976 | 16.537 | 4.515 | medium |
| Contrast $\gamma = 4$ | 0.00% | 0.045 | 0.971 | 15.943 | 4.527 | medium |
| Contrast $\gamma = 8$ | 0.00% | 0.059 | 0.952 | 13.845 | 4.700 | hard |
| Zoom out small | 0.00% | 0.052 | 0.965 | 14.456 | 6.098 | medium |
| Zoom out medium | 0.25% | 0.057 | 0.945 | 14.087 | 5.974 | medium |
| Zoom out large | 3.00% | 0.063 | 0.945 | 13.378 | 6.322 | hard |
| Zoom out extreme | 47.25% | 0.077 | 0.888 | 12.022 | 6.341 | hard |
| Perspective transformation small | 0.00% | 0.053 | 0.964 | 14.382 | 4.664 | medium |
| Perspective transformation medium | 0.00% | 0.055 | 0.962 | 14.149 | 4.705 | medium |
| Perspective transformation large | 0.00% | 0.060 | 0.954 | 13.447 | 4.819 | hard |
| Scale down 1/2 | 0.00% | 0.042 | 0.975 | 16.239 | 4.234 | easy |
| Scale down 1/4 | 0.00% | 0.053 | 0.964 | 13.999 | 4.174 | medium |
| Scale down 1/8 | 0.25% | 0.110 | 0.846 | 7.703 | 4.371 | hard |
| Scale down 1/16 | 17.75% | 0.254 | 0.290 | -0.759 | 6.354 | hard |
| Scale down 1/32 | 100.00% | — | — | — | — | hard |
| Rotation 5° | 0.00% | 0.056 | 0.961 | 13.988 | 4.271 | medium |
| Rotation 15° | 0.00% | 0.069 | 0.944 | 12.056 | 4.387 | medium |
| Rotation 25° | 1.50% | 0.084 | 0.913 | 10.220 | 4.490 | hard |
| Rotation 35° | 5.25% | 0.107 | 0.850 | 8.760 | 4.649 | hard |
| High-frequency noise small | 0.00% | 0.035 | 0.984 | 17.482 | 5.057 | medium |
| High-frequency noise medium | 0.00% | 0.044 | 0.973 | 15.166 | 5.623 | medium |
| High-frequency noise large | 2.00% | 0.058 | 0.963 | 13.054 | 6.079 | hard |
| Low-frequency noise small | 0.00% | 0.038 | 0.985 | 19.420 | 4.842 | medium |
| Low-frequency noise medium | 0.00% | 0.044 | 0.984 | 21.225 | 5.174 | medium |
| Low-frequency noise large | 1.83% | 0.077 | 0.967 | 22.058 | 5.933 | hard |
| Lead opacity 60% | 0.00% | 0.037 | 0.981 | 17.796 | 4.915 | medium |
| Lead opacity 40% | 3.00% | 0.039 | 0.979 | 17.428 | 4.902 | medium |
| Lead opacity 20% | 13.00% | 0.056 | 0.951 | 15.277 | 5.425 | hard |
| Lead opacity 15% | 31.00% | 0.061 | 0.949 | 14.923 | 5.647 | hard |
| Scans | 0.00% | 0.043 | 0.976 | 16.548 | 4.425 | easy |
| Photos Samsung | 0.00% | 0.042 | 0.976 | 16.596 | 4.496 | easy |
| Photos iPhone | 0.00% | 0.041 | 0.978 | 16.647 | 4.398 | easy |
| Photos Doogee | 0.00% | 0.042 | 0.977 | 16.390 | 4.414 | medium |
| Screens | 0.00% | 0.045 | 0.973 | 15.725 | 4.537 | medium |
| Bent paper | 0.00% | 0.049 | 0.959 | 15.176 | 4.603 | medium |
| Crumpled paper | 0.00% | 0.068 | 0.940 | 11.922 | 4.698 | medium |

### ECG digitization timing

The timing of ECG digitization varied, reflecting the impact of image quality and conditions on signal processing. On average, the digitization time across all scenarios was 4.86 seconds (±0.6s), ranging from 3.88 seconds to 6.35 seconds. Low-complexity image subsets, such as those with minimal image compression and optimal brightness, required less processing time, averaging around 4.5 seconds (±0.15s). In contrast, high-complexity subsets representing extreme conditions, such as severe blurring and significant down-scaling, demonstrated longer processing times, with the maximum observed being 6.35 seconds (significantly different from the high-quality cases, P=0.003). Despite these variations, the average time to ECG digitization was consistently under 7 seconds across all scenarios.

### Visual inspection of digitized ECG

In certain cases, the digitization process produced results that appeared nearly perfect upon visual inspection, yet the corresponding performance metrics indicated suboptimal outcomes. For example, as illustrated in Figure 10, an ECG trace that was visually confirmed to be accurate exhibited poor performance metrics in lead V2 (PCC 0.676, RMSE 0.389, and SNR 2.57). Lead V2 exhibits lower metrics due to signal artifacts towards the end of the recording. This anomaly results in null values (NaNs), which, when filled with zeros, contribute to the discrepancy between the observed and expected signal patterns. Lastly, as shown in Figure 11, pacemaker spikes present a significant challenge in the digitization of ECG data. Our digitization solution currently lacks the capability to accurately handle these spikes, resulting in potential inaccuracies in the recorded ECG signals.

**Figure 10.**
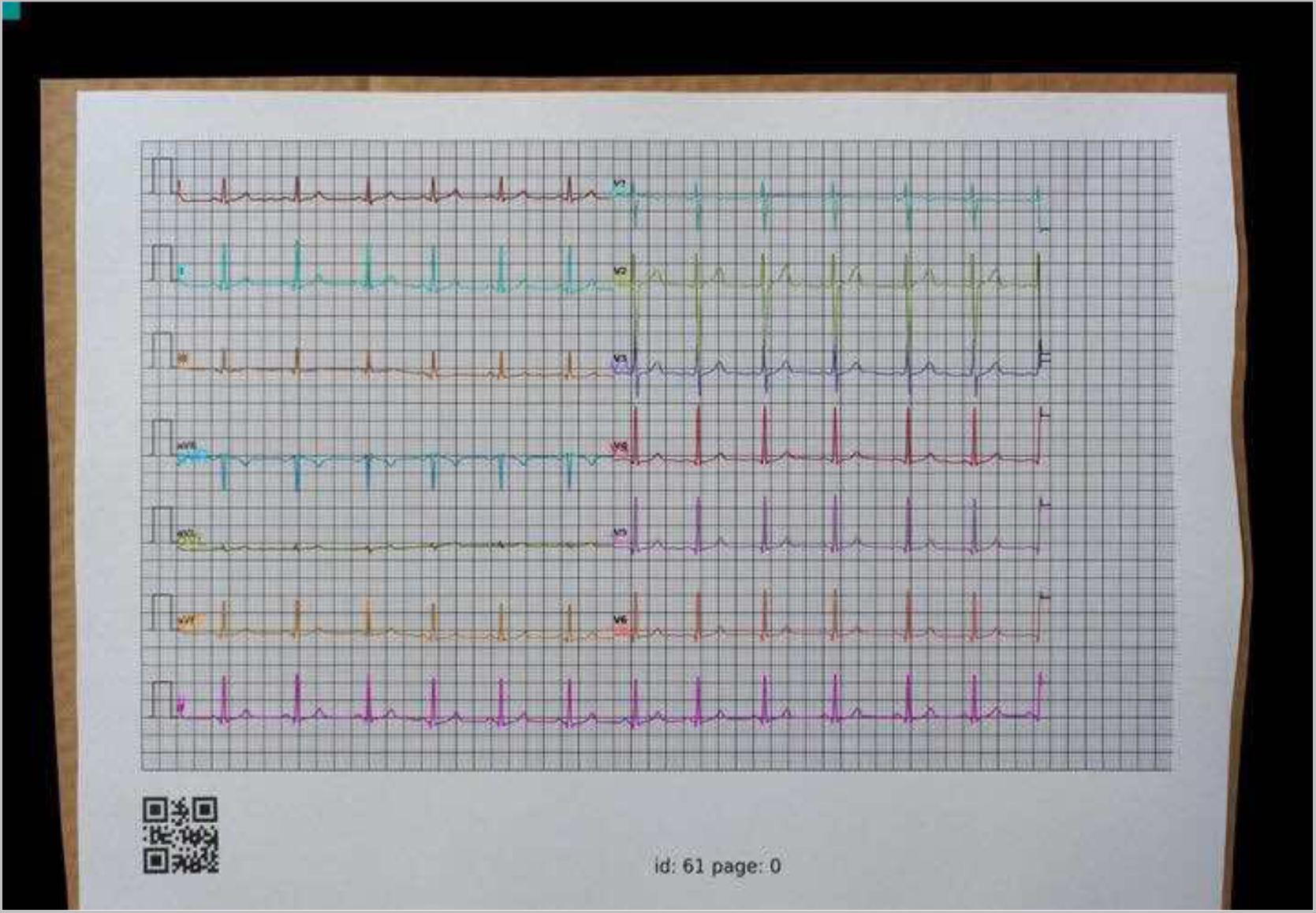
ECG trace of V2 lead, highlighting the problematic section towards the end of the recording. The trace shows deviations from expected patterns, affecting the calculated metrics. Specific metrics for V2: target count – 2500, predicted count – 2431, RMSE – 0.389, PCC – 0.676, and SNR – 2.57.

**Figure 11.**
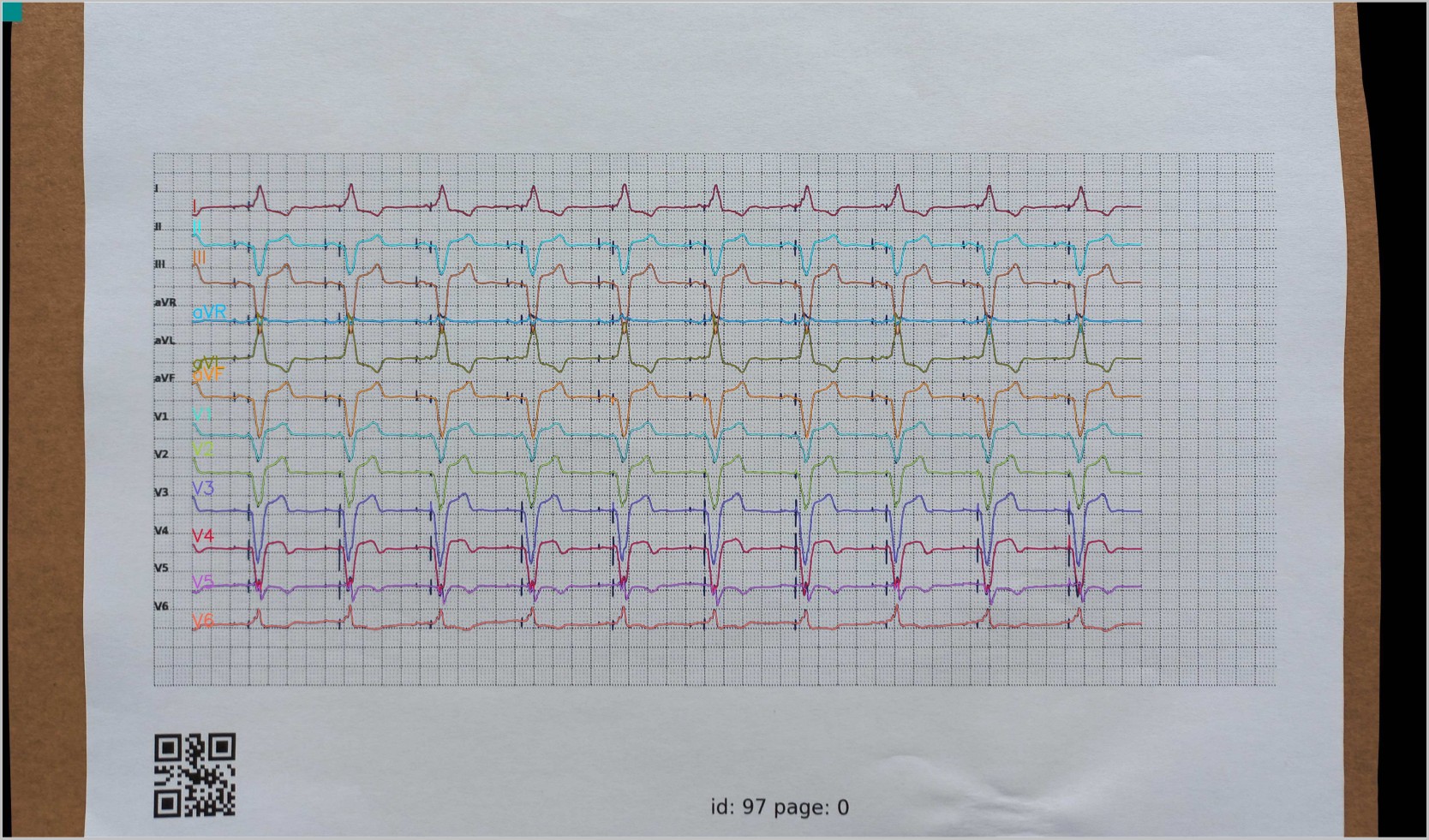
Illustration of ECG traces with visible pacemaker spikes. The digitization solution’s current limitation in handling these spikes is evident, as it fails to differentiate the artificial pacemaker signal from the natural heart rhythm, leading to potential errors in signal interpretation.

## Discussion

This study presents a fully automated, deep learning-based approach to ECG digitization, validated on a large, independent database of 6,000 unique ECG images collected across 60 real-world scenarios. Our approach achieved high performance across individual leads and ECG image scenarios. The average PCC consistently exceeded 91% across all leads with an average RMSE below 0.10 mV and performance decline observed only in extreme image conditions. This level of robustness is essential, given the variability of real-world clinical environments where ECG images are typically captured.

In the context of ECG digitization, several studies have sought to improve the accuracy and reliability of converting image-based ECGs into digital waveforms. Our solution builds upon the advancements made by earlier works while also addressing some of the persistent challenges highlighted in the literature. Traditional approaches, such as those by Lobodzinski et al.^20^ and Badilini et al.^7^, have utilized the underlying millimeter ECG grid to correct skew by identifying and adjusting the skew angle based on selected points on the gridlines. While these methods laid the groundwork for accurate signal extraction, they often required manual intervention and were limited to high-quality scanned images. Badilini et al.^7^ developed ECGScan, a tool designed to convert paper ECGs into digital formats by detecting the grid and extrapolating waveforms using active contour modeling. While the tool demonstrated strong agreement between original and digitized, particularly in terms of waveform reconstruction and QT interval measurements, ECGScan was semi-automated and relied on user input for setting anchor points, limiting its scalability and efficiency. Building on this, Wu et al.^8^ highlighted the need for a fully automated solution and demonstrated the potential of deep learning to automate the entire ECG digitization process with improved accuracy. Moreover, Santamónica et al.^9^ developed an algorithm that incorporates a graphical user interface, yielding a strong correlation. Still, most existing solutions are limited to high-quality images, not fully automatic, and fail to handle the diverse ECG layouts and real-world conditions.

Our approach is characterized by several methodological strengths. First, it is fully automated, and our ECG digitization solution is the first to introduce automatic format detection, eliminating the need to manually specify the ECG layout, image rotation, or lead order, surpassing existing solutions in efficiency and ease of use. Next, we have demonstrated the strength of our solution on a large independent PM-ECG-ID database consisting of a diverse range of real-world image scenarios typically encountered in clinical environments. Unlike previous digitization tools^10^, which often fail in scenarios such as sub-optimal lighting, low-resolution images, and significant image distortions, our method maintains a high level of precision. Even under extreme scenarios such as significant overlapping waveforms, blurring, high levels of image brightness adjustment, and substantial zoom-out, our solution demonstrated robust accuracy, with failure rates remaining low in all but the most complex cases. Moreover, studies^9,10^ on ECG digitization solutions tend to overestimate true performance by discarding cases recorded in extreme conditions or those where their digitization fails. Our paper presents transparent results of our solution and comprehensively evaluates the performance.

Our study also considers the broader clinical implementation of ECG digitization in the context of cardiovascular patient management. Fast acquisition of digitized, de-identified ECG waveforms (consistently under 7 seconds across various image scenarios) allows for rapid and compliant transmission of standardized ECG waveforms in environments where digital integrations are not available. As highlighted by Lence et al.^10^, digitization of ECG images is crucial for advanced ECG interpretation using deep learning algorithms, potentially leading to improved detection. Our approach, with its ability to accurately digitize ECG images captured by smartphones, can enable faster advanced AI ECG interpretation^1,2^ in the non-cardiologist setting.

One of the persistent challenges in ECG digitization has been the accurate processing of overlapping signals, particularly in cases where multiple leads intersect. This is also reflected in our results, which show slightly lower PCC and higher RMSE values in V3-V5. Typically signals with larger amplitudes, while rich in diagnostic information, present greater challenges during the digitization process due to overlap with adjacent leads. These findings are consistent with Wu et al.^8^, reporting a decreased correlation to 60–70% in the same leads due to overlapping lead signals. Likewise, ECGMiner^9^, a tool designed for automated digitization, reported a rejection rate of 14% for the PTB-XL ECG dataset, primarily due to its inability to separate leads that overlap significantly. Nonetheless, the overall performance of our digitization approach remains high across all leads, with an average failure rate of only 6.6%.

## Limitations

Despite the advancements our study presents, there are limitations that warrant further research. Our digitization solution has a significant limitation in accurately digitizing pacing spikes on the ECG. Due to their brief duration and sharp morphology, they pose a unique challenge and are currently not supported in our solution. The PM-ECG-ID, while comprehensive, may not capture the full spectrum of ECG variations encountered in broader clinical practice. Future research should focus on validating our approach across more diverse patient populations and clinical environments to ensure its generalizability. Additionally, extreme image degradation or signal distortion still present challenges that require further algorithm refinement or methods to automatically reject input on the basis of image quality.

Another critical area for future work is the absence of benchmarking datasets in the field of ECG digitization. As noted by Lence et al.^10^, the lack of standardized publicly available datasets makes it difficult to compare the effectiveness of different ECG digitization methods. Our study contributes a publicly available, diverse dataset, which could serve as a model for future studies aiming to establish benchmarks in the field.

Lastly, the performance metrics used to evaluate ECG digitization solutions, such as PCC, RMSE, and SNR, have inherent limitations and often do not accurately reflect clinical relevancy. In several cases, the results of these metrics are discordant with visual inspection of the digitized ECG waveform (Figure 10). For example, a missing P wave in the digitized waveform can have favorable performance metrics, but would severely impact clinical interpretation and patient care. This suggests that while the metrics are useful for standardizing comparisons across different conditions, they may not always align with the clinical usability of the digitized output. It also highlights the inherent limitations of relying solely on quantitative metrics to evaluate the effectiveness of ECG digitization. Potentially, automated segmentation of the digitized ECG signal into clinically relevant waveform segments - such as P waves, QRS complexes, and T waves - followed by the calculation of performance metrics for each of these could provide a more accurate and clinically meaningful assessment of digitization quality.

## Conclusion

In conclusion, our fully automated deep learning-based approach offers high-precision ECG digitization, effectively overcoming the challenges posed by real-world image capture. Our method significantly enhances the accessibility and furthers the utility of ECG data in modern cardiac care, facilitating the adoption of AI-driven interpretation. By enabling rapid and reliable digitization of paper-based ECGs, our approach has the potential to broaden the scope of digital health solutions, improving diagnostic accuracy and patient outcomes across diverse healthcare settings. Future work should focus on further validation in clinical environments and exploring its scalability for widespread adoption.

## Data Availability

All data produced are available online at: https://doi.org/10.5281/zenodo.13617672

## Acknowledgements

The authors would like to thank the clinical experts, study team, data scientists, and AI engineers supporting the data collection, processing, and validation.

## Supplement materials

**Table 1.** ECG digitization compatibility settings.

| Feature | Description |
| --- | --- |
| File Formats | JPEG and PNG |
| Channels | RGB, ensuring consistency in color recognition |
| Voltage Gains | 2.5, 5, 10, and 20 mm/mV |
| Paper Speeds | 5, 12.5, 25, and 50 mm/s |
| Rotation Flexibility | Accepts photos oriented at 0, 90, 180, or 270 degrees |
| Orientational Variations | Handles slight rotations up to 25 degrees |
| Grid Patterns | Supports mm-grid ECGs with 1mm x 1mm squares (minor grid) and thicker borders every 5mm (major grid) |
| Grid-Background Contrast | Variety of paper-grid color combinations, with the grid as the darker element. |
| Electrical Interferences | Detects noisy leads or pacemaker signals |
| Source of Image | Analyzes both scanned versions and photos from screens |
| Physical Imperfections | Manages paper crumples, bends, and slight perspective distortions |
| Optical Challenges | Accommodates different levels of blur, contrast, brightness, and zoom |
| Lead Clarity | Recognizes and processes ECGs with partially missing or faded leads |

**Table 2.**
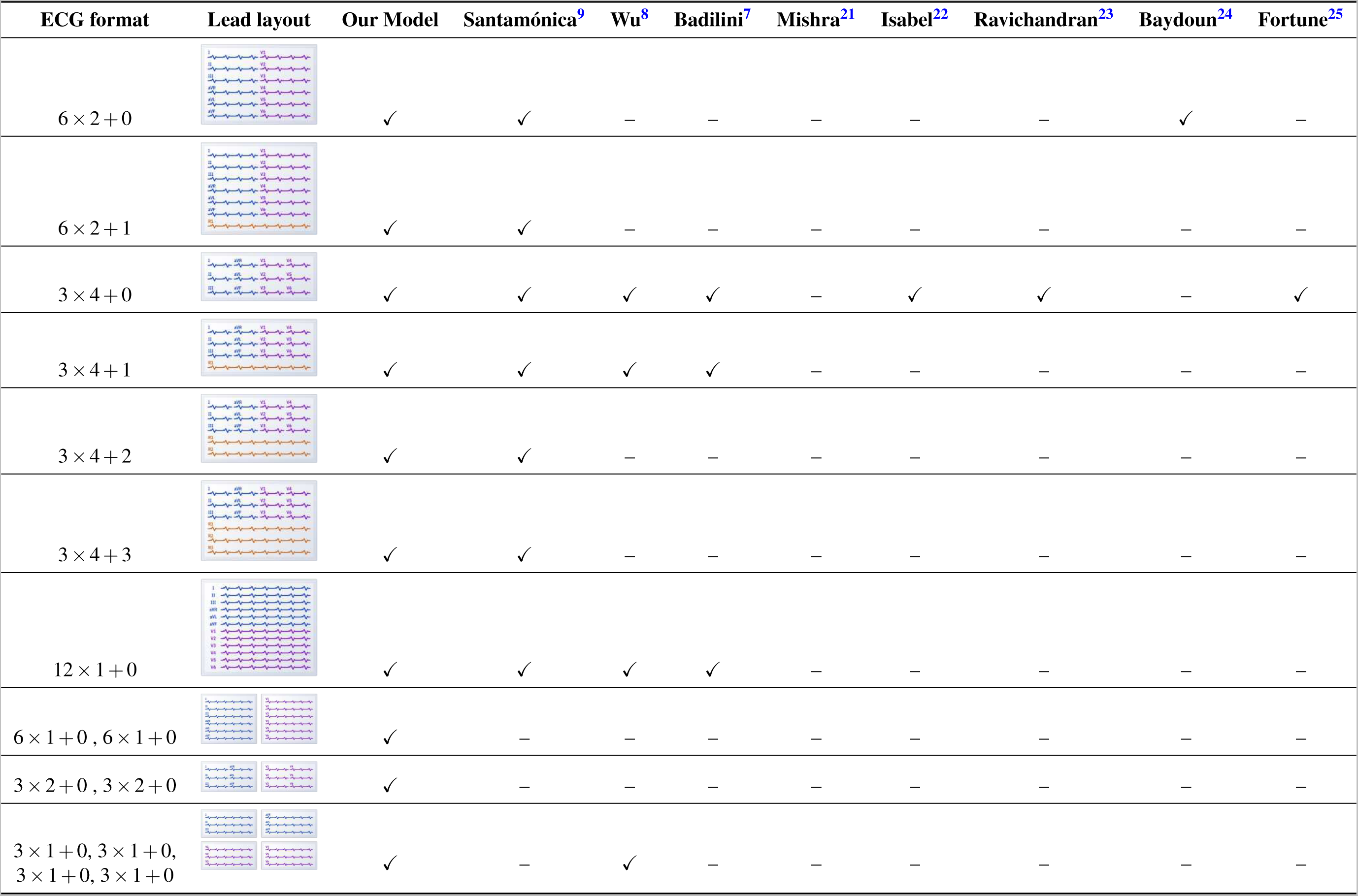
Supported ECG lead layouts. Comparison of different ECG digitization tools: Existing digitization tools are specific ECG configurations or do not detect ECG anchor points by automated methods. By contrast, the automated tool presented here can digitize ECGs of any configuration. (‘—’ suggests that it is unclear if the algorithm can process ECG configuration).

| ECG format | Lead layout | Our Model | Santamónica<sup>9</sup> | Wu<sup>8</sup> | Badilini<sup>7</sup> | Mishra<sup>21</sup> | Isabel<sup>22</sup> | Ravichandran<sup>23</sup> | Baydoun<sup>24</sup> | Fortune<sup>25</sup> |
| --- | --- | --- | --- | --- | --- | --- | --- | --- | --- | --- |
| $6 \times 2 + 0$ | | ✓ | ✓ | — | — | — | — | — | ✓ | — |
| $6 \times 2 + 1$ | | ✓ | ✓ | — | — | — | — | — | — | — |
| $3 \times 4 + 0$ | | ✓ | ✓ | ✓ | ✓ | — | ✓ | ✓ | — | ✓ |
| $3 \times 4 + 1$ | | ✓ | ✓ | ✓ | ✓ | — | — | — | — | — |
| $3 \times 4 + 2$ | | ✓ | ✓ | — | — | — | — | — | — | — |
| $3 \times 4 + 3$ | | ✓ | ✓ | — | — | — | — | — | — | — |
| $12 \times 1 + 0$ | | ✓ | ✓ | ✓ | ✓ | — | — | — | — | — |
| $6 \times 1 + 0, 6 \times 1 + 0$ | | ✓ | — | — | — | — | — | — | — | — |
| $3 \times 2 + 0, 3 \times 2 + 0$ | | ✓ | — | — | — | — | — | — | — | — |
| $3 \times 1 + 0, 3 \times 1 + 0,$<br>$3 \times 1 + 0, 3 \times 1 + 0$ | | ✓ | — | ✓ | — | — | — | — | — | — |

**Table 3.** Metrics for Evaluating ECG Digitization Quality.

| Metric | Description |
| --- | --- |
| **Root Mean Squared Error (RMSE)** | Root Mean Squared Error (RMSE) is the square root of the Mean Squared Error (MSE), quantifying the average of the squares of the differences between predicted (digitized) and actual (original) ECG values. RMSE provides error magnitude in the same units as the ECG data, simplifying clinical interpretation. Lower values indicate higher accuracy, with RMSE being a positive value; the closer to 0, the better. |
| **Pearson Correlation Coefficient (PCC)** | Pearson Correlation Coefficient (PCC) measures the linear correlation between the predicted and actual ECG values. PCC is indicative of the strength of the linear relationship and the predictive accuracy of the digitization process. It ranges from $[-1, 1]$, with 1 indicating perfect positive correlation, -1 indicating perfect negative correlation, and 0 indicating no correlation. |
| **Signal-to-Noise Ratio (SNR)** | Signal-to-Noise Ratio (SNR) compares the level of the desired signal to the level of background noise. SNR is expressed in decibels (dB), with higher values indicating a better quality of the digitized signal. |

**Table 4.** Summary of lead-specific and overall performance metrics for 6,000 ECG entries (100 unique ECGs with 60 augmentations per ECG). The table evaluates four key parameters: average fail rate (%), RMSE, Pearson Correlation Coefficient (PCC), and Signal-to-Noise Ratio (SNR), including cases filled with zeros for failures.

| Lead | Average fail rate | RMSE (zero-filled) | PCC (zero-filled) | SNR (zero-filled) |
| --- | --- | --- | --- | --- |
| **I** | 6.63% | 0.043 | 0.890 | 14.540 |
| **II** | 6.65% | 0.050 | 0.893 | 14.671 |
| **III** | 6.65% | 0.054 | 0.893 | 14.714 |
| **aVR** | 6.62% | 0.038 | 0.887 | 14.224 |
| **aVL** | 6.62% | 0.042 | 0.892 | 14.424 |
| **aVF** | 6.62% | 0.045 | 0.894 | 14.716 |
| **V1** | 6.57% | 0.057 | 0.895 | 15.365 |
| **V2** | 6.57% | 0.096 | 0.887 | 14.125 |
| **V3** | 6.57% | 0.111 | 0.874 | 12.893 |
| **V4** | 6.67% | 0.113 | 0.856 | 11.907 |
| **V5** | 6.67% | 0.094 | 0.873 | 12.585 |
| **V6** | 6.65% | 0.063 | 0.889 | 14.474 |
| **All leads** | 6.62% | 0.067 | 0.885 | 14.053 |

**Table 5.** Comprehensive Evaluation of ECG Digitization Quality Across Varied Conditions: This table details the performance metrics of ECG digitization processes across different datasets subjected to various conditions such as compression, brightness adjustment, and digital noise. Each dataset contains 100 ECGs and is assessed based on Root Mean Square Error (RMSE), Pearson Correlation Coefficient (PCC), and Signal-to-Noise Ratio (SNR), including failures filled with zeros.

| Dataset | failed | RMSE<br>(zero-filled) | PCC<br>(zero-filled) | SNR<br>(zero-filled) | Average time [s] | Quality group |
| --- | --- | --- | --- | --- | --- | --- |
| **Digital image original** | **0.00%** | **0.037** | **0.981** | **17.644** | **4.915** | **easy** |
| JPEG compression 90% | 0.00% | 0.041 | 0.977 | 16.608 | 4.593 | easy |
| JPEG compression 45% | 0.00% | 0.042 | 0.976 | 16.526 | 4.509 | easy |
| JPEG compression 22% | 0.00% | 0.042 | 0.976 | 16.473 | 4.504 | medium |
| JPEG compression 11% | 0.00% | 0.044 | 0.974 | 16.148 | 4.510 | medium |
| JPEG compression 5% | 0.00% | 0.049 | 0.968 | 15.258 | 4.507 | hard |
| JPEG compression 2% | 0.00% | 0.064 | 0.945 | 13.449 | 4.456 | hard |
| Blur kernel 11 | 0.00% | 0.046 | 0.972 | 15.771 | 4.567 | medium |
| Blur kernel 21 | 1.00% | 0.061 | 0.947 | 13.310 | 5.391 | medium |
| Blur kernel 31 | 69.00% | 0.234 | 0.113 | -0.036 | 6.128 | hard |
| Blur kernel 41 | 96.00% | 0.232 | 0.003 | -0.022 | 3.883 | hard |
| Brightness -40 | 0.00% | 0.041 | 0.977 | 16.581 | 4.577 | easy |
| Brightness -80 | 0.00% | 0.043 | 0.974 | 16.292 | 4.539 | medium |
| Brightness -120 | 0.00% | 0.046 | 0.971 | 15.676 | 4.451 | medium |
| Brightness -160 | 5.25% | 0.080 | 0.868 | 11.586 | 5.056 | hard |
| Brightness +40 | 0.00% | 0.041 | 0.977 | 16.669 | 4.525 | easy |
| Brightness +80 | 0.00% | 0.042 | 0.976 | 16.603 | 4.566 | medium |
| Brightness +120 | 0.00% | 0.042 | 0.976 | 16.446 | 4.641 | medium |
| Brightness +160 | 0.00% | 0.048 | 0.963 | 15.617 | 4.679 | hard |
| Contrast $\gamma = 0.125$ | 0.00% | 0.040 | 0.977 | 16.688 | 4.633 | medium |
| Contrast $\gamma = 0.25$ | 0.00% | 0.039 | 0.978 | 16.909 | 4.567 | medium |
| Contrast $\gamma = 0.5$ | 0.00% | 0.040 | 0.977 | 16.823 | 4.537 | easy |
| Contrast $\gamma = 0.666$ | 0.00% | 0.040 | 0.977 | 16.757 | 4.561 | easy |
| Contrast $\gamma = 1.5$ | 0.00% | 0.042 | 0.976 | 16.543 | 4.580 | easy |
| Contrast $\gamma = 2$ | 0.00% | 0.041 | 0.976 | 16.537 | 4.515 | medium |
| Contrast $\gamma = 4$ | 0.00% | 0.045 | 0.971 | 15.943 | 4.527 | medium |
| Contrast $\gamma = 8$ | 0.00% | 0.059 | 0.952 | 13.845 | 4.700 | hard |
| Zoom out small | 0.00% | 0.052 | 0.965 | 14.456 | 6.098 | medium |
| Zoom out medium | 0.25% | 0.058 | 0.943 | 14.051 | 5.974 | medium |
| Zoom out large | 3.00% | 0.067 | 0.917 | 12.974 | 6.322 | hard |
| Zoom out extreme | 47.25% | 0.147 | 0.468 | 6.343 | 6.341 | hard |
| Perspective transformation small | 0.00% | 0.053 | 0.964 | 14.382 | 4.664 | medium |
| Perspective transformation medium | 0.00% | 0.055 | 0.962 | 14.149 | 4.705 | medium |
| Perspective transformation large | 0.00% | 0.060 | 0.954 | 13.447 | 4.819 | hard |
| Scale down 1/2 | 0.00% | 0.042 | 0.975 | 16.239 | 4.234 | easy |
| Scale down 1/4 | 0.00% | 0.053 | 0.964 | 13.999 | 4.174 | medium |
| Scale down 1/8 | 0.25% | 0.110 | 0.844 | 7.686 | 4.371 | hard |
| Scale down 1/16 | 17.75% | 0.245 | 0.238 | -0.624 | 6.354 | hard |
| Scale down 1/32 | 100.00% | 0.231 | 0.000 | -0.000 | – | hard |
| Rotation 5° | 0.00% | 0.056 | 0.961 | 13.988 | 4.271 | medium |
| Rotation 15° | 0.00% | 0.069 | 0.944 | 12.056 | 4.387 | medium |
| Rotation 25° | 1.50% | 0.085 | 0.899 | 10.064 | 4.490 | hard |
| Rotation 35° | 5.25% | 0.113 | 0.806 | 8.303 | 4.649 | hard |
| High-frequency noise small | 0.00% | 0.035 | 0.984 | 17.482 | 5.057 | medium |
| High-frequency noise medium | 0.00% | 0.044 | 0.973 | 15.166 | 5.623 | medium |
| High-frequency noise large | 2.00% | 0.060 | 0.943 | 13.054 | 6.079 | hard |
| Low-frequency noise small | 0.00% | 0.038 | 0.985 | 19.420 | 4.842 | medium |
| Low-frequency noise medium | 0.00% | 0.044 | 0.984 | 21.225 | 5.174 | medium |
| Low-frequency noise large | 1.83% | 0.084 | 0.949 | 21.655 | 5.933 | hard |
| Lead opacity 60% | 0.00% | 0.037 | 0.981 | 17.796 | 4.915 | medium |
| Lead opacity 40% | 3.00% | 0.044 | 0.950 | 16.905 | 4.902 | medium |
| Lead opacity 20% | 13.00% | 0.077 | 0.828 | 13.291 | 5.425 | hard |
| Lead opacity 15% | 31.00% | 0.109 | 0.654 | 10.297 | 5.647 | hard |
| Scans | 0.00% | 0.043 | 0.976 | 16.548 | 4.425 | easy |
| Photos Samsung | 0.00% | 0.042 | 0.976 | 16.596 | 4.496 | easy |
| Photos iPhone | 0.00% | 0.041 | 0.978 | 16.647 | 4.398 | easy |
| Photos Doogee | 0.00% | 0.042 | 0.977 | 16.390 | 4.414 | medium |
| Screens | 0.00% | 0.045 | 0.973 | 15.725 | 4.537 | medium |
| Bent paper | 0.00% | 0.049 | 0.959 | 15.176 | 4.603 | medium |
| Crumpled paper | 0.00% | 0.068 | 0.940 | 11.922 | 4.698 | medium |

## Appendix 1 ECG printout variations and image augmentations

- **Base photos**: These printouts were captured under ideal lighting and positioning conditions, providing a high-quality baseline for comparison with other conditions. The device used for taking these base photos was the Samsung Galaxy A32, which features a 64.2 MP main camera with an aperture of f/1.8.
- **Medium distance photos**: Printouts were captured at varying distances to simulate different real-world scenarios and to provide additional variations in perspective. Medium-distance photos were taken with the camera positioned approximately 35 cm away from the ECG, simulating scenarios where the photo was taken from a slight distance.
- **Longer distance photos**: Longer distance photos were captured with the camera positioned around 53 cm from the ECG, allowing for further variation in perspective and enabling the application of different zoom levels and rotations during post-processing.
- **Photos using different phones**: Printouts were captured using two smartphone models to account for differences in camera quality and image processing algorithms:

**–** iPhone 11 with a 12 MP main camera and an aperture of f/1.8

**–** Doogee S55 with a 13 MP main camera and an aperture of f/2.0

Despite the Doogee S55’s higher resolution camera, the iPhone 11’s overall image quality surpasses it due to several factors. The iPhone 11’s advanced aperture (f/1.8), superior Image Signal Processor (ISP), larger sensor, and Optical Image Stabilization enable it to deliver superior light gathering, color analysis, low light performance, and image stability.

- **Scanned printouts**: All ECGs in this category were scanned using a single scanner model in color and at a resolution of 300 DPI.
- **Screen photos**: Photos were captured by photographing ECGs displayed on a Philips screen, which exhibited the Moire effect.
- **Photos of printouts with bends**: The conditions simulate real-world scenarios where the ECG printout may be held in one hand and a photo is taken with the other hand, causing the printout to crease or bend during the process or by using a printout with a bend that could have been caused by folding.
- **Photos of printouts with crumples**: ECG photos were captured after intentionally folding and crumpling the printout, mimicking situations where an ECG printout is folded up for storage and later unfolded, potentially causing the paper to be crumpled or damaged in the process.

To enhance the diversity of the database and better simulate real-world challenges, we applied a variety of augmentations to the base photos, distance photos, and digital ECG waveforms. The digital ECG waveforms with applied augmentations were visualized as JPEG images with a resolution of 7, 483 ⇥ 5, 291 pixels.

For the **base photos**, we manually introduced the following augmentations:

- **Contrast adjustment**: We adjusted the contrast of the photos using gamma correction to simulate varying lighting conditions and to challenge algorithms to recognize ECG patterns with suboptimal contrast. We applied gamma correction with the following unitless gamma values: {0.125, 0.25, 0.5, 0.667, 1.5, 2, 4, 8}. These values nonlinearly control the contrast transformation curve, with lower gamma values (*<* 1) resulting in brighter photos with increased contrast and higher gamma values (*>* 1) leading to darker photos with decreased contrast.
- **Brightness modification**: We manipulated the brightness of photos by transforming them into the HSV color space and modifying only the brightness channel. The brightness values were adjusted within a range of 0-255, with 0 representing complete darkness and 255 representing maximum brightness. To achieve this, we applied a set of adjustments: {±40, ±80, ±120, ±160}. This process simulates situations where photos may be overexposed or underexposed due to external lighting conditions or camera settings. The adjustments influence the pixel values of the photos, with positive values increasing brightness and negative values decreasing it.
- **Photo blurring**: We applied varying levels of blur to the photos, representing scenarios where photos might be captured with slight motion blur or out-of-focus due to camera shake or incorrect focus settings. We used blur kernels of the following sizes in pixels: {11, 21, 31, 41}. Smaller kernel sizes (e.g., 11) result in a mild blurring effect, while larger kernel sizes (e.g., 41) produce a more significant blurring effect. The kernel size determines the degree of blurring by controlling the area over which the image pixels are averaged or smoothed, with larger kernels leading to more extensive averaging and greater blur.
- **JPEG compression**: We introduced JPEG compression to simulate photo quality degradation caused by file compression and transmission, testing the resilience of algorithms against lossy photo formats. The compression was applied at various levels, represented by JPEG quality factors: {90, 45, 22, 11, 5, 2}. For instance, a quality factor of 90 denotes minimal compression and, consequently, relatively superior preservation of photo quality. Conversely, a quality factor of 2 corresponds to substantial compression, leading to significantly diminished photo quality. It is crucial to note that these quality factors do not directly imply the resultant file size as a percentage of the original size, as the relationship between the quality factor and file size is non-linear and content-dependent. Therefore, these figures primarily serve to reflect the extent of photo quality retention or degradation.
- **Resolution alteration**: We changed the resolution of the photos to represent various display and capture resolutions, ensuring that the algorithms can perform well across different device specifications and display sizes. We reduced the resolution by the power of 2, which involves halving the sides of the photo in each step, effectively reducing the overall photo size (width x height) by a square of the factor used. The following factors were used: {2, 4, 8, 16, 32}. For example, a reduction by a factor of 2 results in a photo with sides half the length of the original (1/4 size), while a reduction by a factor of 4 results in a photo with sides one-fourth the length of the original (1/16 size). This process simulates the impact of reduced photo resolution on the performance of digitization algorithms across a range of resolutions commonly encountered in real-world scenarios.

All the utilized visualizations are demonstrated in Figure 1a-1e. For the **distance photos**, we applied the following augmentations, as shown in Figure 1f-1h:

- **Perspective transformation**: We applied perspective transformations to simulate different camera angles, which can occur when users take photos of ECGs from various positions, testing the algorithms’ ability to recognize patterns under such conditions. We assumed an 80-degree angle of view for a Samsung camera and calculated the distance to the photo accordingly. We then rotated the photo plane around the *x* and *y* axes, such that the sum of the two angles was equal to {20, 30, 40} degrees, and projected the transformed photo onto the focal plane. Rotations around the *z*-axis are covered in a separate "Rotation" augmentation, ensuring that the database includes a comprehensive representation of potential camera rotations and angles that may be encountered in real-world scenarios.
- **Rotation**: We rotated the photos by varying degrees to represent possible photo orientations, ensuring that the algorithms can recognize ECG patterns even when photos are not perfectly aligned. The photos were rotated by {5, 15, 25, 35} degrees.
- **ECG-to-padding ratio adjustment**: We adjusted the ECG-to-padding ratio of the photos to capture varying levels of detail, challenging the algorithms to identify ECG patterns across different scales. Using two sets of ECG photos with surrounding padding, we have created datasets with different levels of surrounding padding. These levels are represented by the following categories: {small, medium, large, extra_large}.

For the selected **ECG waveforms**, we manually applied the following augmentations:

- **Lead visibility modification**: We modified the lead visibility by introducing four additional alpha levels, representing intermediate and worse visibility conditions, simulating cases where the leads may be partially obscured or faintly printed on the ECG paper. We changed the opacity in Matplotlib using the following values: {1, 0.6, 0.4, 0.2, 0.15}, where 1 represents full opacity (no modification) and lower values represent decreased lead visibility due to increased transparency.
- **Introduction of different frequency noises**: We generated new ECG waveforms with baseline wandering and muscle frequency interference by applying various frequency noises. This augmentation tests the ability of the algorithms to digitize ECGs accurately, even in the presence of noise. In real-world applications, ECGs can be affected by baseline wandering and other types of noise, making it challenging to extract accurate ECG patterns. Therefore, this augmentation is essential to assess the robustness of the digitization algorithms to noise artifacts. We introduced two types of noise:

**–** High-frequency noise (frequency range: 20-40 Hz) simulates muscle frequency interference, which can be caused by muscle contractions or other physiological processes. We applied three levels of amplitude to represent varying levels of interference:

* Low: amplitude range 0.02-0.05 mV

* Middle: amplitude range 0.05-0.1 mV

* Large: amplitude range 0.1-0.15 mV

**–** Low-frequency noise (simulating baseline wanderings, frequency range: 0.05-0.3 Hz) represents fluctuations in the ECG baseline due to factors such as respiration or body movement. We introduced three levels of amplitude to represent varying degrees of baseline wandering:

* Low: amplitude range 0.1-0.25 mV

* Middle: amplitude range 0.25-0.5 mV

* Large: amplitude range 0.5-0.75 mV

Examples of the visualization of ECG waveforms with applied augmentations are depicted in Figure 2.

As detailed in Table 1, our comprehensive and diverse set of augmentation techniques, ranging from different photo variations to waveform transformations, enabled the assembly of an extensive ECG image database with 6,000 unique entries, all designed to rigorously test and improve ECG digitization algorithms.

**Figure 1.**
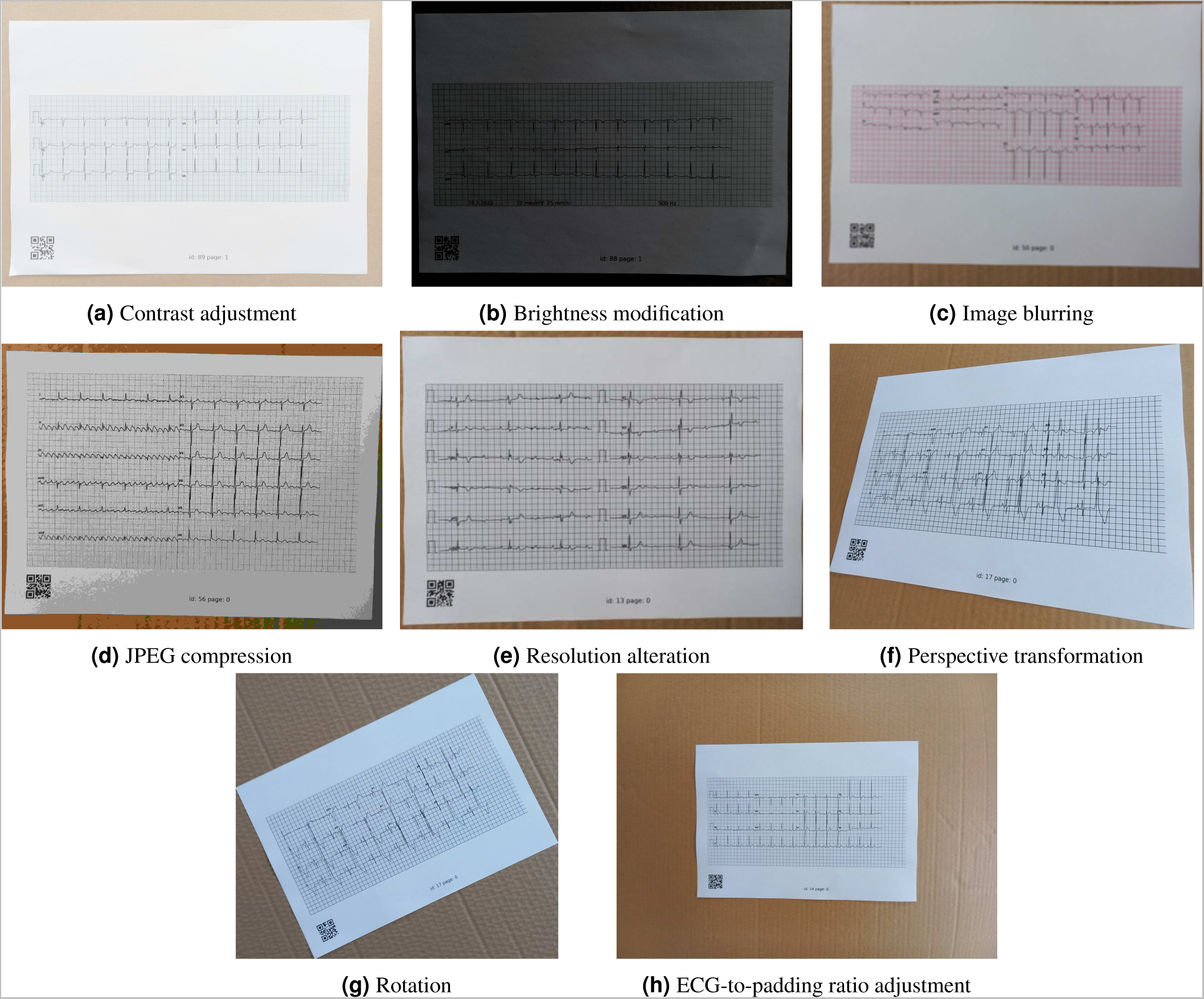
Examples of base/distance photos with used augmentations.

**Figure 2.**
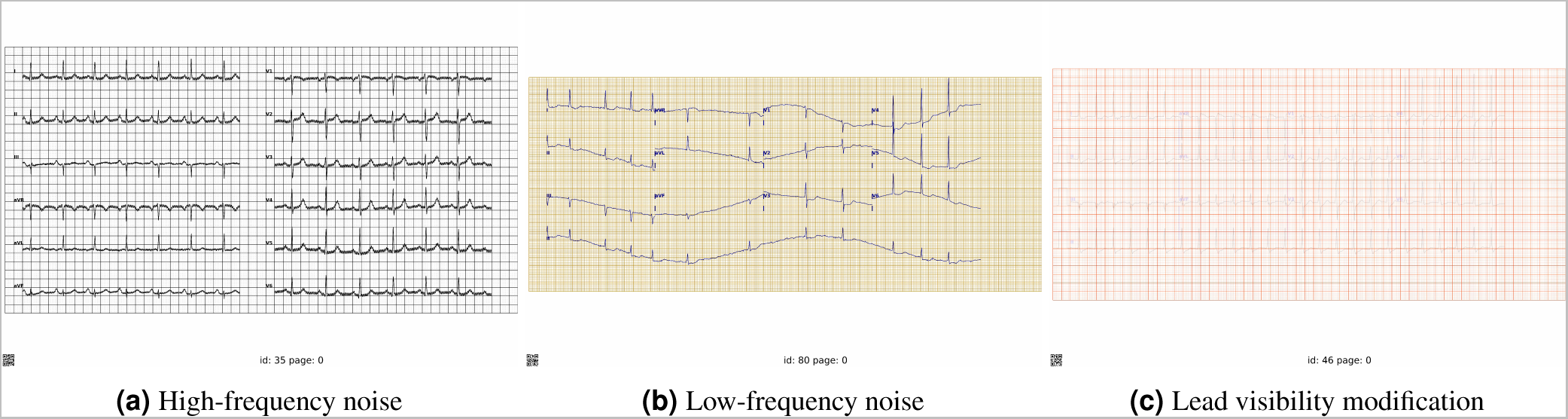
Examples of visualization of ECG waveforms with used augmentations.

**Table 1.** Detailed distribution of ECGs across various categories and methodologies.

| Category | Methodology | Number of ECGs |
| --- | --- | --- |
| Photo variations | Photos with bends | $1 \times 100$ |
| | Photos with crumples | $1 \times 100$ |
| | Photos from iPhone | $1 \times 100$ |
| | Photos from Doogee | $1 \times 100$ |
| | Photos from Samsung | $1 \times 100$ |
| | Scanned printouts | $1 \times 100$ |
| | Screen photos | $1 \times 100$ |
| Samsung photo augmentations | Image blurring | $4 \times 100$ |
| | Brightness modification | $8 \times 100$ |
| | Contrast adjustment | $8 \times 100$ |
| | JPEG compression | $6 \times 100$ |
| | Resolution alteration | $5 \times 100$ |
| Distance photo augmentations | ECG-to-padding ratio adjustment | $4 \times 100$ |
| | Perspective transformation | $3 \times 100$ |
| | Rotation | $4 \times 100$ |
| Waveform transformations | High-frequency noise | $3 \times 100$ |
| | Low-frequency noise | $3 \times 100$ |
| | Lead visibility modification | $5 \times 100$ |
| **Total** |  | **6,000** |

